## Supplementary Figures for "mRNA COVID-19 vaccine booster fosters B and T cell responses in immunocompromised patients"

### Online supplemental material

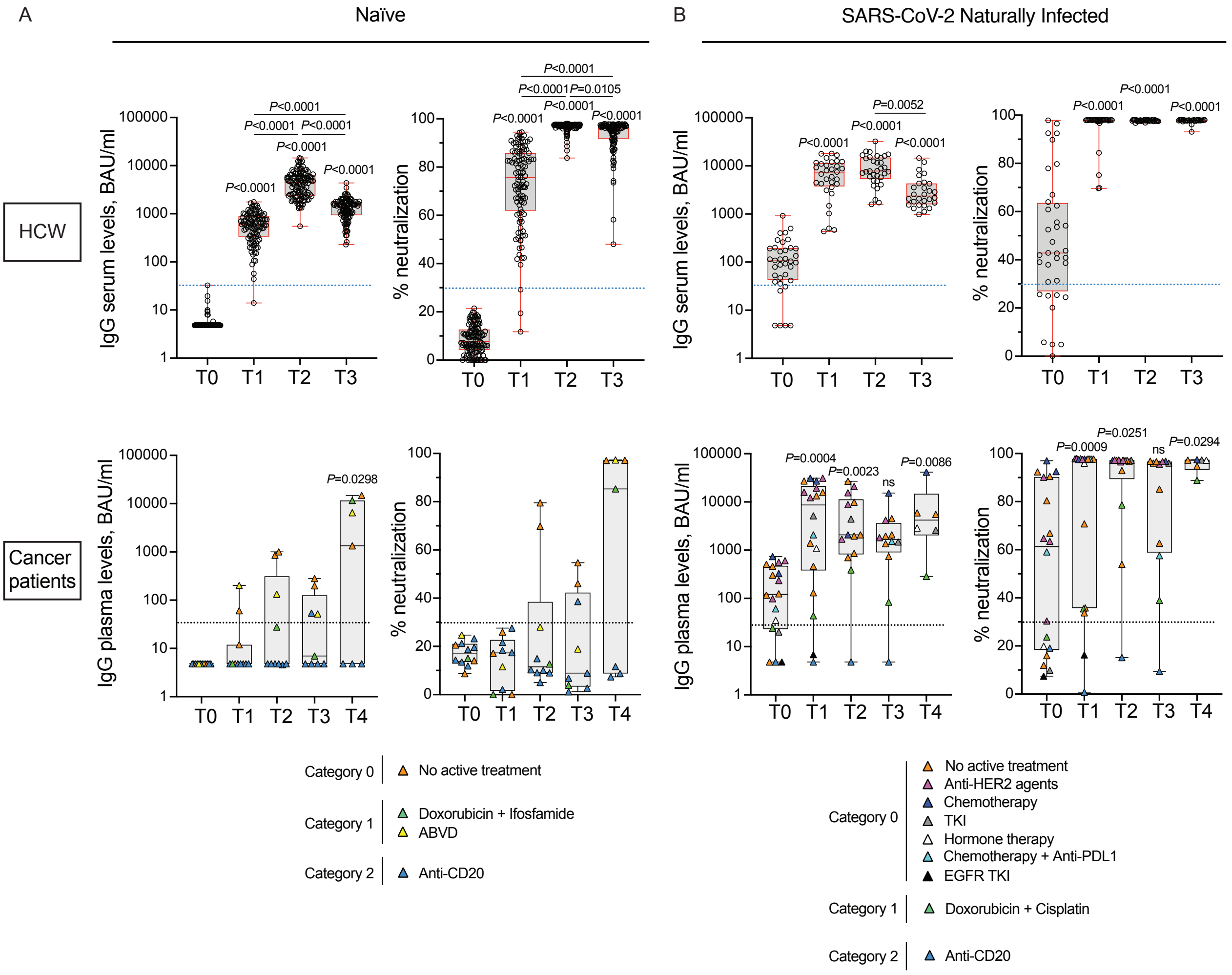


**Figure S1. SARS-CoV-2 naïve cancer patients treated with anti-CD20 fail to produce neutralizing antibodies.**

IgG antibody response and its neutralizing activity were measured in serum of vaccinated naïve (HCW, n=125; Cancer patients, n=12) (A) and SARS-CoV-2 naturally infected (HCW, n=36; Cancer patients, n=18) (B) health care workers (HCW) and cancer patients at different time points (T0, T1, T2, T3). Cancer patients received also the booster dose and sera were analyzed at 2 weeks after the third dose (T4, naïve, n=7; SARS-CoV-2 naturally infected, n=6). Samples ≥ 33.8 BAU/mL (IgG plasma levels) or ≥ 30% signal inhibition (neutralization) were considered positive (dotted blue and black lines). For IgG serum levels, log scale on y axis. The box plots show the interquartile range, the horizontal lines show the median values, and the whiskers indicate the minimum-to-maximum range. Each dot corresponds to an individual subject. *P* values were determined using 2-tailed Kruskal-Wallis test with Dunn's multiple comparisons post test. *P* values refer to baseline (T0) when there are no connecting lines. Cancer patients were classified according to the type of treatment: no active treatment or low (category 0, orange), medium (category 1, green) or high (category 2, blue) interference with the immune system. The distribution of patients in each category and the type of treatment are indicated in the legend.

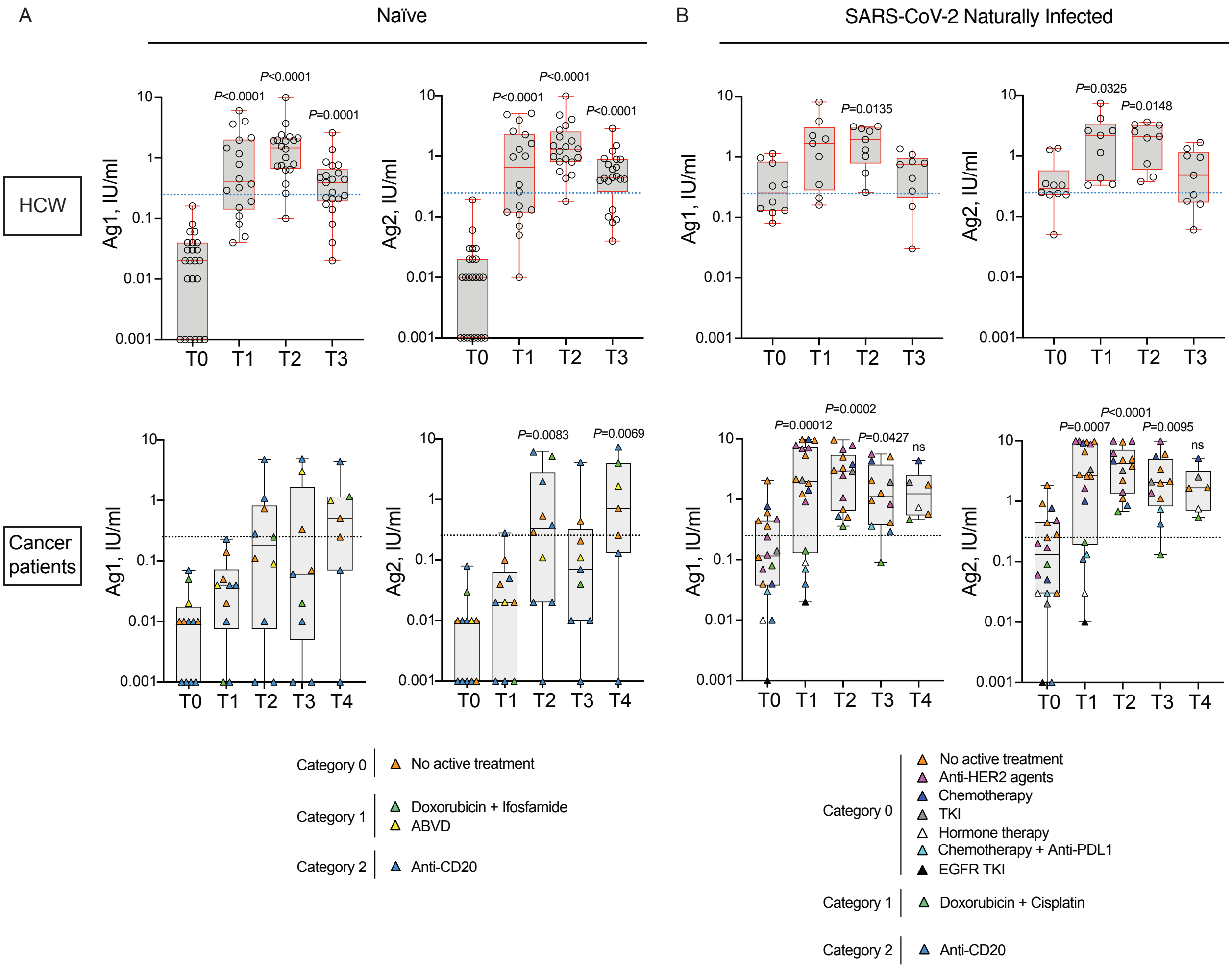


**Figure S2. SARS-CoV-2 naïve cancer patients treated with anti-CD20 may fail to activate T cell responses.**

Anti-spike T cell response activation, by using specific CD4 (Ag1) and CD4 plus CD8 (Ag2) T cell epitopes of the spike protein, were measured in plasma of vaccinated naïve (HCW, n=24; Cancer patients, n=12) (A) and SARS-CoV-2 naturally infected (HCW, n=10; Cancer patients, n=18) (B) health care workers (HCW) and cancer patients at different time points (T0, T1, T2, T3). Cancer patients received also the booster dose and plasma were analyzed at 2 weeks after the third dose (T4, naïve, n=7; SARS-CoV-2 naturally infected, n=6). Samples ≥ 0.25 IU/mL were considered positive (dotted blue and black lines). Log scale on y axis. The box plots show the interquartile range, the horizontal lines show the median values, and the whiskers indicate the minimum-to-maximum range. Each dot corresponds to an individual subject. *P* values were determined using 2-tailed Kruskal-Wallis test with Dunn's multiple comparisons post test. *P* values refer to baseline (T0) when there are no connecting lines. Cancer patients were classified according to the type of treatment: no active treatment or low (category 0, orange), medium (category 1, green) or high (category 2, blue) interference with the immune system. The distribution of patients in each category and the type of treatment are indicated in the legend.

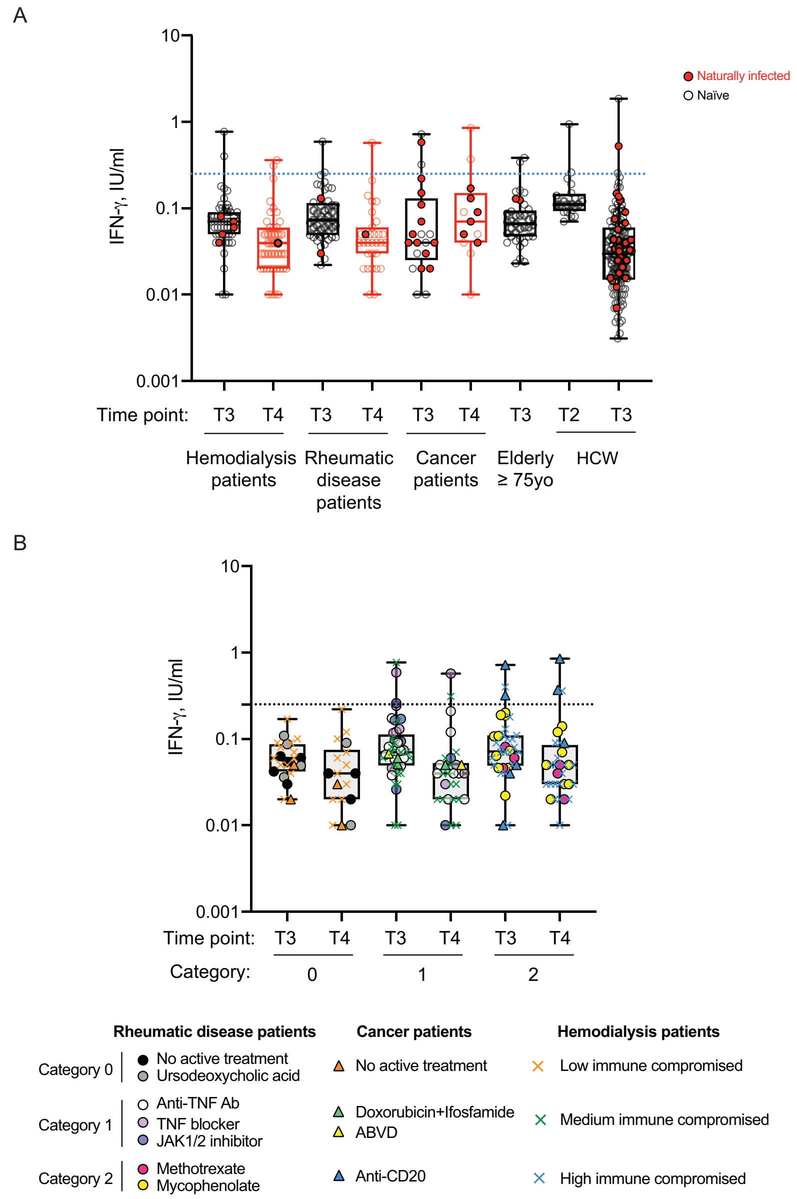


**Figure S3. The IFN-γ basal levels in naïve and SARS-CoV-2 naturally infected vaccinated subjects.**
(A) IFN-γ basal level measured in naïve vaccinated (white circles) and SARS-CoV-2 naturally infected (red circles) health care workers (HCW, n=132), elderly people ≥75 yo (n=37), cancer patients (n=21), patients with rheumatic diseases (n=48) or in patients in hemodialysis (n=53) subjects at different time points.
(B) IFN-γ basal level measured in naïve vaccinated immunocompromised patients with cancer (n=9), rheumatic diseases (n=47) or patients in hemodialysis (n=48) at 2-4 months after second dose (T3) and 2 weeks after the booster dose (T4). Cancer and rheumatic disease patients were classified according to the type of treatment: no active treatment or low (category 0), medium (category 1) or high (category 2) interference with the immune system, whereas patients in hemodialysis were classified with an immunoscore related to the disease for which the patients are in dialysis and their comorbidities: low (category 0), medium (category 1) or high (category 2) immune compromised. Samples ≥ 0.25 IU/mL were considered positive (dotted black lines). The distribution of patients in each category and the type of treatment are indicated in the legend.

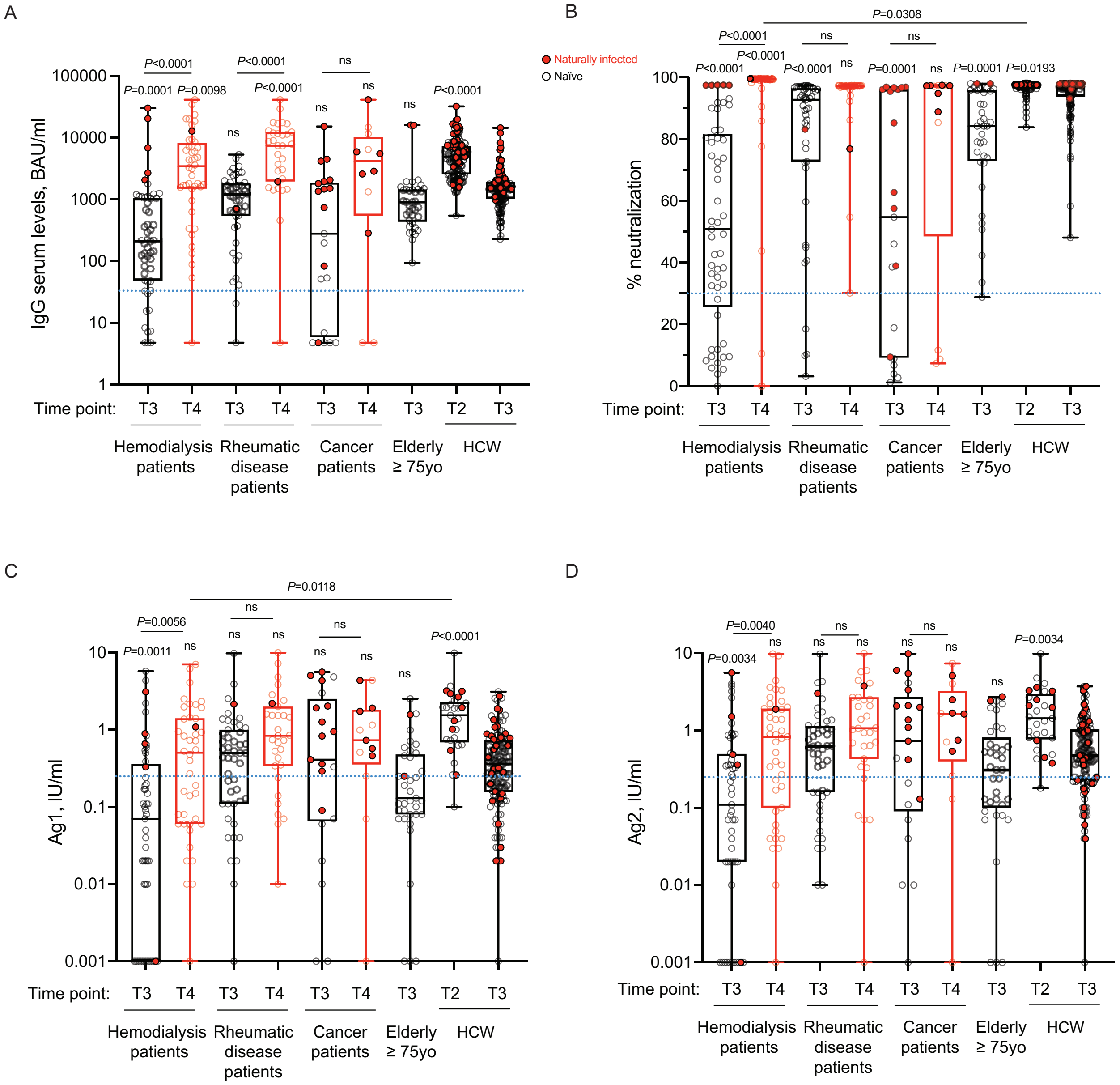


**Figure S4. The immune response in naïve and SARS-CoV-2 naturally infected immunocompromised patients at 2-4 months after the second dose and 2 weeks after the booster dose.**

IgG antibody response (A), its neutralizing activity (B) and anti-spike T cell response activation, by using specific CD4 (Ag1, C) and CD4 plus CD8 (Ag2, D) T cell epitopes of the spike protein were measured in serum and plasma of vaccinated naïve (white circles) and SARS-CoV-2 naturally infected (red circles) health care workers (HCW, n=132), elderly people ≥75 yo (n=37), cancer patients (n=21), patients with rheumatic diseases (n=48) or in patients in hemodialysis (n=53) at 2-4 months after second dose (black, T3) or at 2 weeks after the third dose (red, T4). As a control, we indicated values of IgGs, their neutralizing activity and anti-spike T cell response activation of vaccinated naïve and SARS-CoV-2 naturally infected health care workers (HCW, n=152) at 10 days after the second dose (T2). The box plots show the interquartile range, the horizontal lines show the median values, and the whiskers indicate the minimum-to-maximum range. Each dot corresponds to an individual subject. P values were determined using 2-tailed Kruskal-Wallis test with Dunn's multiple comparisons post test. P values refer to HCW T3 when there are no connecting lines. Positivity was based on: anti-spike IgG ≥ 33.8 BAU/mL; neutralization ≥ 30% and T cell response ≥ 0.25 IU/mL for either Ag1 or Ag2 (dotted blue lines).

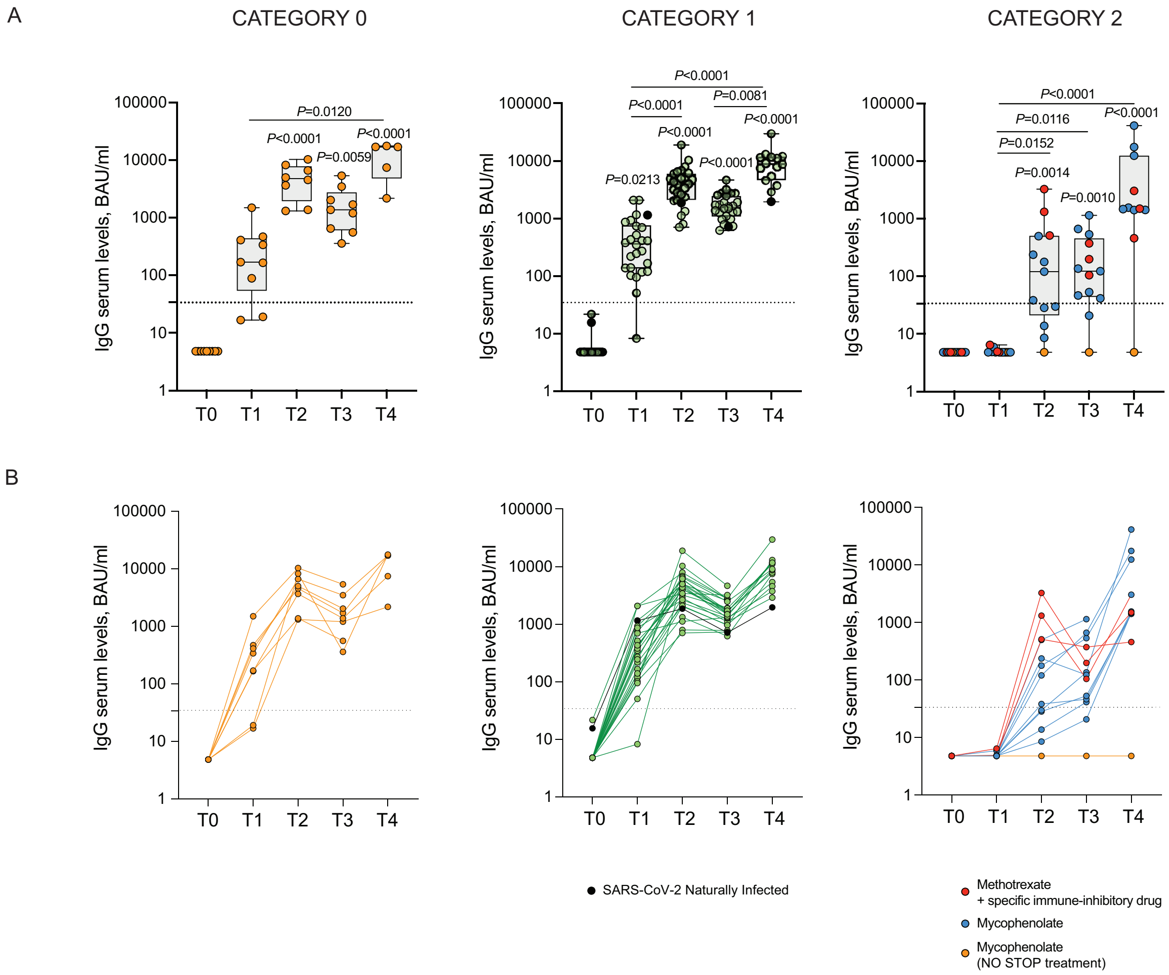


**Figure S5. Kinetics of antibody response in rheumatic disease patients according to treatment.** IgG antibody response was measured in serum of vaccinated patients whit rheumatic diseases at different time points (T0, T1, T2, T3, T4). Patients were classified according to the type of treatment (category 0, n=9; 1, n=26; 2, n=13). Samples ≥ 33.8 BAU/mL were considered positive (dotted black lines). Log scale on y axis. (A) The box plots show the interquartile range, the horizontal lines show the median values, and the whiskers indicate the minimum-to-maximum range. Each dot corresponds to an individual subject. (B) Spaghetti plots showing the trends for each individual subject by linked dots. P values were determined using 2-tailed Kruskal-Wallis test with Dunn's multiple comparisons test (A). P values refer to baseline (T0) when there are no connecting lines.

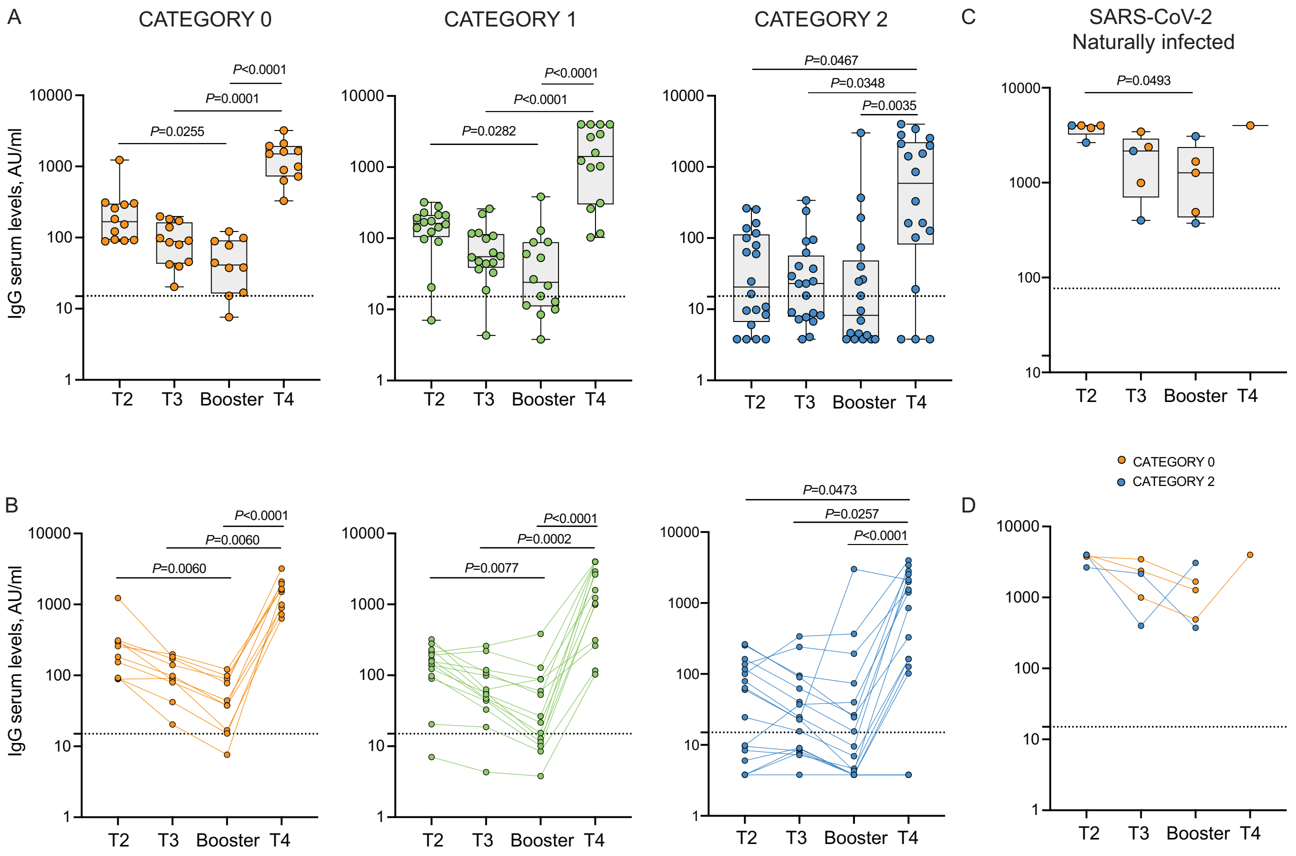


**Figure S6. Kinetics of antibody response in patients in hemodialysis according to treatment.** IgG antibody response was measured in serum of vaccinated naïve (A, B) and SARS-CoV-2 Naturally Infected (C, D) patients in hemodialysis at different time points (T2, T3, at the time of the booster dose – 6 months from the second dose – and T4). Patients were classified with an immunoscore related to the disease for which the patients are in dialysis and their comorbidities (category 0, n=12 naïve and n=3 SARS-CoV-2 Naturally Infected; category 1, n=16; category 2, n=20 naïve and n=2 SARS-CoV-2 Naturally Infected). Samples ≥ 15 AU/mL were considered positive (dotted black lines). Log scale on y axis. (A, C) The box plots show the interquartile range, the horizontal lines show the median values, and the whiskers indicate the minimum-to-maximum range. Each dot corresponds to an individual subject. (B, D) Spaghetti plots showing the trends for each individual subject by linked dots. P values were determined using 2-tailed Kruskal-Wallis test with Dunn's multiple comparisons test (A, C) or Friedman test with Dunn's multiple comparisons test (B). P values refer to baseline (T0) when there are no connecting lines.
